# Development of Entrustable Professional Activities for the University of New Mexico Nephrology Fellowship Training Program

**DOI:** 10.64898/2026.01.05.26343456

**Authors:** Huzefa Y Saria, Hayley P Israel, J. Pedro Teixeira, Namita Singh, Christos P Argyropoulos, Sara A Combs, Maria-Eleni Roumelioti

## Abstract

**Rationale & Objective:** Competency-based medical education emphasizes observable skills rather than time-based training. Entrustable Professional Activities (EPA) transform competencies into distinct, assessable clinical tasks but have not yet been systematically developed for U.S nephrology fellowships. We aimed to create and achieve consensus on a set of nephrology-specific EPAs and align them with Accreditation Council for Graduate Medical Education (ACGME) competency standards.

**Study Design:** A consensus framework was developed using an online 3-round modified Delphi method.

**Settings & Participants:** Study was conducted within the University of New Mexico nephrology fellowship program. Participants included eight faculty nephrologists and one nephrology fellow.

**Analytical Approach:** An initial EPA list was generated by the study team using program objectives, literature review, and clinician insight. Participants rated each EPA using a 5-point Likert scale with consensus requiring strict criteria. Final EPAs were independently mapped to ACGME nephrology program requirements to ensure alignment with national competency.

**Results:** Nine study participants (100% response rate) completed all survey rounds. Through iterative consensus, utilizing strict criteria, a final list of 22 distinct EPAs were achieved covering core domains of practice including dialysis management, acute kidney injury, chronic kidney disease, electrolyte abnormalities, hypertension, kidney stones, glomerular disease, pregnancy, transplant care, and education. Mapping demonstrated that the EPAs sufficiently corresponded to the breadth of ACGME-required sub competencies, offering a practical framework for translating broad milestones into observable clinical tasks.

**Limitations:** The study was conducted at a single fellowship program with a small number of participants which may limit generalizability. Implementation feasibility, resource implications, and potential unintended consequences such as checklist mentality and documentation burden were evaluated during a subsequent phase of the study.

**Conclusions:** We developed the first consensus-consensus based set of EPAs geared for U.S based nephrology fellowship programs while being systematically aligned with ACGME program requirements. This framework provides a foundation for standardized assessment and curriculum development in nephrology and may inform broader efforts to implement EPA-based evaluation across fellowship programs nationally.

## Background

Competency Based Medical Education (CBME) has transformed postgraduate medical training by shifting the focus from time-based progression to the demonstration of essential skills and competencies required for independent clinical practice.^1^ CBME is grounded in clearly defined competencies that trainees must acquire, often structured around frameworks such as the Accreditation Council for Graduate Medical Education (ACGME) competencies and milestones. However, while these frameworks establish broad domains of competency, they often lack direct applicability to day-to-day clinical decision-making and real-world practice.^2^ To bridge this gap, Entrustable Professional Activities (EPAs) have emerged as a practical tool within CBME, translating these competencies into specific, observable, and assessable clinical tasks that reflect the actual responsibilities of a physician.

EPAs are formulated units of clinical work that can quantifiably measure the competency of a professional in training. By aligning assessment with workplace-based tasks, EPAs provide a structured approach to evaluating clinical competence, ensuring standardization in training while allowing for individualized learning progression.

The main aspect of EPAs includes the performance of a task by a trainee under direct supervision. The passing of responsibility to a trainee to complete a task has been deemed the entrustment decision.^3^ A supervising physician will entrust a task to a trainee which they deem appropriate for their level of competency. EPAs are uniquely suited for the medical profession as most daily activities of care and treatment that a provider performs can be broken down into self-contained tasks that can be individually tracked. This allows a training program to monitor and assess the competency level of their trainees through quantifiable metrics which they can apply to the broader ACGME competencies.

ACGME Internal Medicine competencies were introduced to fellowship programs in 2014. The fellow is regularly assessed in their progress to perform in 23 sub-competencies within six main competencies, with named milestones within each sub-competency to guide the clinical competency committee in determining where the fellow is performing. The six major competencies are Patient Care, Medical Knowledge, Professionalism, Interpersonal Communication Skills, Practice-Based Learning and Improvement, and Systems-Based Practice.

Currently, the training and competency-based assessment we provide and use in Nephrology fellowship programs can leave gaps in content and clinical knowledge necessary for successful independent practice, especially for those graduates entering non-academic practices. Our graduating fellows must develop skillsets aligned to national practice environments, including patient safety and development of professional practice. EPAs are a tool that programs could utilize to help ensure their fellows are successfully developing these necessary skills. However, there is a lack of consensus regarding EPA development and their use in US Nephrology fellowship programs.^4^

The most recent study on the development of nephrology-specific EPAs comes from Tanaka et al., who applied a modified Delphi method to inform the development of EPAs for IM residents rotating through a nephrology department in a Japanese academic hospital.^4^ While narrower in scope and geared towards a different learner population, Tanaka’s methodology strongly influenced the design of our own study. Specifically, we adopted their strict consensus criteria and the use of modified Delphi method for constructing a consensus list of program-specific EPAs.^4^

Therefore, in this study, we sought to address this unmet need to develop EPAs for Nephrology fellowship training. These EPAs could be applied after site-specific modifications to other programs as well. We first focused on determining learners’ and educators’ perspectives and needs regarding work-based assessment to develop EPAs for our Nephrology training program at the University of New Mexico (UNM). We aimed to develop a list of EPAs that represent the scope of practice in nephrology both nationwide as well as in the state of New Mexico. In addition, we matched the EPAs we developed to the available ACGME competencies to create concrete goals and milestones for fellows.

## Methods

All study forms and procedures were approved by our local IRB (HRPO# 23-338)

### Creation of Initial EPA List

The initial list of EPAs was developed by members of the research group after three online meetings (from October 2023 to April 2024). The group included four experienced attending physicians, one medical education specialist, and two 2^nd^-year nephrology fellows. The initial list was created by referencing and adapting annually reviewed training goals and behavioral objectives of the Division of Nephrology at UNM. Textbook chapter headings and search lists of the most common diagnostic codes were considered. An iterative process was used to develop this initial list, reorganizing after each of several discussions and adhering to the following conditions:^5^

- The EPAs should be part of the essential professional work of nephrology and not general medical ability.
- Must require adequate knowledge, skill, and attitude.
- Must lead to recognized performance that is unique to a doctor and should be unique to physicians in nephrology.
- Should be independently executable within a specific time frame.
- Should be observable and measurable in its process and outcome.
- Should reflect one or more of the ACGME competency categories.

The development of each EPA’s description followed the guidelines set by ten Cate ^6^ (Table S1). Based on review of the literature and the 23 ACGME sub-competencies required for Nephrology training, we hypothesized that we would have to develop at least 10 EPAs to be able to cover most of the various clinical aspects of work of a practicing nephrologist.

### Consensus Development Method

Similar to prior studies which successfully developed EPAs, we used the modified Delphi method as our consensus-building approach.^4^

We found that the modified Delphi approach was particularly well-suited to the design of our study that consisted of a pre-constructed list which was iterated in an asynchronous setting maintaining anonymity within our participants.

### Recruitment of Survey Respondents

After a brief presentation of the project to the nephrology division, potential study participants were approached through a confidential e-mail that included the study’s consent form. For this study we approached attending physicians working both at UNM Hospital (UNMH) and the New Mexico Veterans Affairs (VA) Health Care System since both organizations train fellows in our program. We also approached all 1^st^ and 2^nd^ year fellows except for the two fellows that were study co-investigators. There was a 100% response rate for all surveys sent form the consented study participants.

### Survey

The survey was created by the research group and consisted of two sections, one which collected respondent demographic information and a second focused on the evaluation of each of the proposed EPAs. The emailed survey included a training tool with the scope to explain EPAs and the entrustment decision-making process and to provide participants with instructions on how to complete the survey. The survey respondents were asked to evaluate each proposed EPA on a 5-point Likert scale, ranging from 1 = EPA should definitively be excluded to 5 = EPA should definitively be included.

After each proposed EPA, the respondent could add comments or items that were reviewed for the next cycle/year of the study (implementation and evaluation cycle). Three rounds of survey administration occurred between 4/18/2024 and 5/28/2024.

### Consensus Standard

A discussion among the study investigators took place after each response cycle to finalize which EPAs would be included in the following cycle of refinement. Consensus was achieved when: the mean rating was ≥4 with a standard deviation of <1 and at least 75% of respondents rated the EPA ≥4.^4^ A total of three cycles were performed to reach a final consensus (Table 2).

### Matching the EPAs to the ACGME Milestones

Two independent reviewers (HS and MER) mapped our final list of EPAs onto the *ACGME Program Requirements for Graduate Medical Education in Nephrology*. Mapping the EPAs onto the ACGME requirements allowed us to explore whether the final list of EPAs successfully represented the overall scope of nationally required nephrology training (see Table 3).

## Results

### Survey Respondents

Our study population consisted of 9 survey respondents: 8 faculty nephrologists recruited from both the UNMH and the VA hospital, and 1 nephrology fellow. All attendings and the fellow that initially consented to participate in the study completed the 3 circles of the survey. The collected demographic data for the participant group is displayed in Table 1.

**Table 1:** Participant Group Demographics.

| Parameters |  |
| --- | --- |
| Age (mean ± SD) | 47.2 (±8.7) |
| Females (%) | 44.4 |
| Attendings (%) | 88.9 |
| Fellows (%) | 11.1 |
| Years of clinical experience (median, IQR)<sup>a</sup> | 12.5 (17.5) |

<sup>a</sup>Years of clinical experience included both academic and private practice environments

The initial EPA creation resulted in 25 separate EPAs. The consensus building process led to the removal of EPAs numbered 11, 13, and 14 (Table S2). The final list of EPAs (n=22) including the marked removed EPAs are displayed in Table 2.

**Table 2:** Final List of the Developed EPAs.

| **Final EPA List** |
| --- |
| **Chronic Kidney Disease** |
| 1) Assessing and providing an initial plan for investigation/work-up and management of patients with CKD (reduced eGFR and/or proteinuria). |
| 2) Monitoring and providing medical management and comprehensive care for patients with stable kidney disease or progressive kidney dysfunction. |
| 3) Facilitating patients' transition to an ESKD treatment modality or to end-of-life care, being able to identify eligible patients and to discuss all available therapeutic options including, if applicable, the provision of kidney supportive measures with the patient and/or their caregivers. |
| **Hemodialysis** |
| 4) Ordering and adjusting dialysis prescriptions for uncomplicated patients with ESKD. |
| 5) Assessing and providing initial management for patients with common a) acute (e.g., hypotension, bleeding, cramping, nausea/vomiting, etc.), and b) chronic complications of HD (e.g., itching) and HD access (e.g., thrombosis, stenosis, infections, etc.) |
| 6) Providing longitudinal management for patients receiving chronic HD. |
| **Peritoneal Dialysis** |
| 7) Ordering and adjusting PD prescriptions for uncomplicated patients with ESKD. |
| 8) Assessing and managing the care of patients with a) infectious and b) non-infectious complications of PD. |
| 9) Providing longitudinal management for patients receiving chronic PD. |
| **Kidney Transplantation** |
| 10) Assessing and providing initial management for transplant patients with acute kidney allograft dysfunction. |
| 11) Admitting patients to undergo kidney transplantation. <sup>a</sup> |
| 12) Providing postoperative care for kidney transplant recipients with an uncomplicated or complex course. |
| 13) Assessing and providing initial management for patients with common infectious and non-infectious complications of kidney transplantation. <sup>a</sup> |
| 14) Assessing the eligibility of patients with kidney disease for kidney transplantation. <sup>a</sup> |
| 15) Monitoring patients receiving immune-modulating therapy and managing kidney transplant-related complications. |
| **Acute Kidney Injury** |
| 16) Assessing, establishing, and providing an initial comprehensive management plan for patients with AKI. |
| 17) Ordering and adjusting RRT prescriptions for patients with AKI and other acute/urgent indications for RRT. |
| **Electrolyte and Acid-Base Abnormalities** |
| 18) Assessing and providing an initial investigation and management plan for patients with complex fluid and electrolyte abnormalities. |
| **Hypertension** |
| 19) Management of uncontrolled hypertension in hospitalized patients with CKD assessing their complications and target of treatment. |
| 20) Assessing and treating patients with difficult-to-control idiopathic or suspected secondary hypertension. |
| **Kidney Stones** |
| 21) Initial and longitudinal management plan of patients with kidney stones. |
| **Glomerular Diseases** |
| 22) Initial diagnostic and management plan for patients with nephrotic or nephritic syndrome. |
| 23) Longitudinal management of GN patients receiving immune or non-immune therapy and management of therapy-related complications. |
| **Pregnancy** |
| 24) Integrating knowledge of the effects of pregnancy, pregnancy outcomes, kidney disease, and its treatment in the care of women with kidney disease. |
| **Teaching and Education** |
| 25) Delivering scholarly teaching to a variety of audiences, including peers, junior trainees, and/or other health professionals. |

Abbreviations: CKD; Chronic Kidney Disease, eGFR; estimated Glomerular Filtration Rate, ESKD; End-Stage Kidney Disease, HD; Hemodialysis, PD; Peritoneal Dialysis, AKI; Acute Kidney Injury, RRT; Renal Replacement Therapy, GN; Glomerulonephritis/<sup>a</sup> Indicated EPAs were eliminated throughout the consensus building process

The final list of developed EPAs was matched to the ACGME sub-competency milestones by two independent study members and consensus was reached (Table 3). Throughout the matching process, sub competencies IV.B.1.c).(1).(a), IV.B.1.b).(2).(a).(iii), IV.B.1.b).(2).(b).(iii), and IV.B.1.c).(1).(b) were matched the most with each being matched to five separate EPAs.

**Table 3.** Final EPA List Matched with ACGME Subcompetencies.

| EPA Number<sup>a</sup> | ACGME Subcompetencies<sup>b</sup> |
| --- | --- |
| 1 | IV.B.1.b).(1).(a).(ii),<br>IV.B.1.b).(2).(a).(i),<br>IV.B.1.b).(2).(a).(ii),<br>IV.B.1.b).(2).(b).(iv),<br>IV.B.1.c).(1).(j),<br>IV.B.1.e) |
| 2 | IV.B.1.b).(1).(a).(ii),<br>IV.B.1.b).(2).(a).(ii),<br>IV.B.1.c).(1).(a)<br>IV.B.1.c).(1).(g) |
| 3 | IV.B.1.b).(1).(a).(vi),<br>IV.B.1.b).(2).(a).(iii) |
| 4 | IV.B.1.b).(1).(a).(vi),<br>IV.B.1.b).(2).(a).(iii),<br>IV.B.1.b).(2).(b).(i)<br>IV.B.1.b).(2).(b).(iii),<br>IV.B.1.c).(1).(a),<br>IV.B.1.c).(1).(b),<br>IV.B.1.c).(1).(c),<br>IV.B.1.c).(1).(o),<br>IV.B.1.c).(1).(q) |
| 5 | IV.B.1.b).(2).(b).(i),<br>IV.B.1.c).(1).(b).(v),<br>IV.B.1.c).(1).(b).(vi) |
| 6 | IV.B.1.b).(2).(a).(iii),<br>IV.B.1.b).(2).(b).(i),<br>IV.B.1.b).(2).(b).(iii),<br>IV.B.1.c).(1).(b),<br>IV.B.1.c).(1).(o),<br>IV.B.1.c).(1).(r),<br>IV.B.1.f) |
| 7 | IV.B.1.b).(1).(a).(vi),<br>IV.B.1.b).(2).(a).(iii),<br>IV.B.1.b).(2).(b).(iii)<br>IV.B.1.c).(1).(b),<br>IV.B.1.c).(1).(c),<br>IV.B.1.c).(1).(o),<br>IV.B.1.c).(1).(q) |
| 8 | IV.B.1.b).(2).(b).(iii),<br>IV.B.1.c).(1).(b),<br>IV.B.1.c).(1).(q),<br>IV.B.1.c).(1).(r) |
| **9** | IV.B.1.b).(2).(a).(iii),<br>IV.B.1.b).(2).(b).(iii),<br>IV.B.1.c).(1).(b),<br>IV.B.1.c).(1).(r),<br>IV.B.1.f) |
| **10** | IV.B.1.b).(1).(b),<br>IV.B.1.c).(1).(l).(vii) |
| **12** | IV.B.1.b).(1).(b),<br>IV.B.1.c).(1).(l) |
| **15** | IV.B.1.b).(1).(b),<br>IV.B.1.c).(1).(a),<br>IV.B.1.c).(1).(h),<br>IV.B.1.c).(1).(l) |
| **16** | IV.B.1.b).(1).(a).(i) |
| **17** | IV.B.1.b).(1).(a).(i),<br>IV.B.1.b).(2).(b).(ii),<br>IV.B.1.c).(1).(a),<br>IV.C.5.h) |
| **18** | IV.B.1.b).(1).(a).(iii),<br>IV.B.1.c).(1).(e) |
| **19** | IV.B.1.b).(1).(a).(ii),<br>IV.B.1.b).(1).(a).(x),<br>IV.B.1.c).(1).(d) |
| **20** | IV.B.1.b).(1).(a).(x),<br>IV.B.1.c).(1).(d),<br>IV.B.1.f) |
| **21** | IV.B.1.b).(1).(a).(iv),<br>IV.B.1.c).(1).(f),<br>IV.B.1.c).(1).(i) |
| **22** | IV.B.1.b).(1).(a).(ix),<br>IV.B.1.b).(2).(b).(iv),<br>IV.B.1.c).(1).(g),<br>IV.B.1.c).(1).(h) |
| **23** | IV.B.1.b).(1).(a).(ix),<br>IV.B.1.c).(1).(a),<br>IV.B.1.c).(1).(h) |
| **24** | IV.B.1.b).(1).(a).(xi) |
| **25** | IV.D.1.b),<br>IV.D.2.b) |

<sup>a</sup> EPAs numbered 11, 13, and 14 were not matched due to elimination during consensus building process.
<sup>b</sup> For details regarding numbered competency refer to ACGME Program Requirements for Graduate Medical Education in Nephrology<sup>13</sup>

## Discussion

The ACGME has established a structured competency-based framework for the assessment of trainees in all ACGME-accredited programs, including adult nephrology fellowship.^7,8^ While the ACGME has created these frameworks to evaluate fellows during training, the actual competencies remain broad and abstract in their language, making them challenging to translate into use in practical evaluation. For example, within the Patient Care domain, the sub-competency *Chronic Dialysis Therapy (PC2)*, outlines progressive levels of proficiency.^7^ At the highest level, the milestone describes a fellow’s ability to “[identify] the complexities of providing quality care to a population of patients receiving dialysis” and “[anticipate] and [manage] the breadth of comorbid medical and technical complications in the patient on dialysis, including when dialysis is not appropriate”.^7^ While this may seemingly encompass in a broad sense the necessary skills a nephrology fellow needs to be proficient in the care of patients on chronic dialysis, it lacks specificity on how these skills can and should be demonstrated, observed, or assessed in routine clinical practice. This ultimately creates a gap between regulatory language and the day-to-day supervisory judgments that a program director and supervising physician must make when determining a fellow’s readiness in managing this aspect of patient care. Our study sought to address these limitations by using the framework of EPAs to create observable and trackable units of clinical work that reflect the most essential responsibilities a practicing nephrologist must be able to perform independently regardless of their site of practice at the completion of their fellowship.

Although CBME has gained increasing traction in postgraduate training across multiple specialties, the implementation of structured assessment tools in the field of nephrology remains limited and highly variable.^9^ Several efforts over the past decade have made strides in addressing this gap by providing foundational work in milestone development, curricular mapping, and EPA development. One of the earliest calls for nephrology-centered competency structures came from a proposal published by Yuan et al. in 2013.^10^ They advocated for milestones that are both quantitatively measurable and objectively defined to reduce both evaluator bias and improve reproducibility among programs.^10^ They also emphasized the value of simplicity and parsimony in evaluation metrics, meaning that a higher number of milestones would not equate to better outcomes. Rather, simpler and succinctly defined milestones would lead to better tracking, evaluation, and outcomes for the program.^10^

Our EPA development efforts directly echo several recommendations made by Yuan et al. such as their call for simplicity and objectively defined metrics. For example, if we recall the ACGME nephrology milestone PC2, our equivalent EPAs that cover the patient care aspect of chronic dialysis are addressed by EPAs 4,5,6 for hemodialysis and 7,8,9 for peritoneal dialysis. While the ACGME milestone uses a generalized statement to cover all aspects of chronic dialysis treatment, our six EPAs each descriptively call for evaluation of a specific aspect of chronic dialysis care. EPA 4, for example, addresses ordering and managing prescriptions of patients on hemodialysis while EPA 5 focuses on managing the complications of hemodialysis. These EPAs can be more readily linked to skill performance.

Subsequent work by the same authors further explores the operationalization of nephrology-centered milestones and provides a framework for mapping ACGME’s initial 23 IM sub-competency milestones to a nephrology schema.^11^ They describe a limited set of five EPAs and associate each with suggested tools for assessment. However, two main limitations apply to this initial framework. The study primarily focused on curricular mapping and descriptive categorization of nephrology-based competencies. Secondly, the developed EPAs were not formulated through a structured consensus process and limited in scope, addressing only five aspects of nephrology practice. In contrast, our study centered around the goal of developing a widely generalizable set of relatively comprehensive nephrology-specific EPAs that were derived from expert agreement and aggregated through strict consensus criteria. Furthermore, our study began with principle clinical proficiencies and worked to match each to preexisting ACGME core competencies and educational program requirements. This approach grounded our EPA list not only in educational theory but in practice-based consensus.

Outside the U.S., efforts to redefine nephrology training assessment with EPA-based models have also emerged. In Canada, the Royal College of Physicians and Surgeons launched the Competence by Design (CBD) initiative to transition postgraduate training from a time-based to an outcomes-based structure.^12^ As a part of this national reform, nephrology-specific EPAs were developed for both adult and pediatric settings. These EPAs were categorized across four stages of training and were each mapped to CanMEDS milestone domains.^12^ Our study parallels this work but leans on ACGME competency standards to be widely applicable in U.S postgraduate training. Another point of distinction is that, while the Canadian framework was led by a central regulatory body with national authority for implementation, our EPA development reflects a grassroots consensus from U.S.-based nephrology educators and program leaders working at a fellowship that often trains fellows destined for both academic and community-based careers. Our study highlights an important first step in aligning national fellowship expectations in nephrology with an EPA-based framework grounded in U.S. based practice realities.

We chose to utilize the *ACGME Program Requirements for Graduate Medical Education in Nephrology* as the primary resource for milestones for matching our created EPAs rather than the *ACGME’s Nephrology Subspecialty Milestones*.^7,13^ While the subspecialty milestones serve as critical developmental benchmarks for assessing fellow progression, they continue to remain broad and difficult to translate into discrete clinical tasks. The accreditation requirements provide more structured and detailed sets of competencies for various domains of nephrology training that can be matched to our EPAs. By grounding our EPA framework in program accreditation standards, we ensure that our EPAs not only perform their function as observable and assessable units of training but also align with national expectations for fellowship curricula.

This study offers several notable strengths. First, the EPA development process was grounded in consensus from an expert panel that included both nephrology faculty and nephrology fellows. This ensured that the resulting EPAs reflected not only the supervisory expectations of the faculty but also the lived educational realities of trainees. Second, participants were recruited from both UNMH and the affiliated VA Medical Center, allowing us to integrate insights from nephrologists into two distinct practice settings serving distinct patient populations.

However, the study is not without its limitations. Most significantly, despite our goal to make it widely applicable, the EPA development process was conducted at a single fellowship program which could limit the generalizability of the EPA framework to other programs across the U.S. Future multicenter studies should aim to validate these EPAs nationally to build consensus on the adaptability of these developed EPAs. Moreover, we are cognizant of various conceptual and practical pitfalls that come with the implementation of competency-based education frameworks. One concern is that the implementation of EPAs may increase documentation burden for both faculty and learners. Indeed, the implementation of an EPA framework comes with additional logistical burden on programs which individual institutions will need to account for. Our current study does not address the infrastructure or resources required to implement EPA-based assessment frameworks, but this should be the subject of future research. Recognizing these limitations underscores the importance of future research aimed at refining these EPAs and determining optimal approaches to EPA implementation.

## Conclusions and Future Directions

This study represents a first step in establishing a structured, consensus-driven EPA framework tailored to nephrology fellowship training in the U.S. By aligning each EPA with ACGME accreditation requirements, milestones, and core competencies, we aimed to develop a set of observable and assessable tasks that encompass the most crucial aspects of nephrology fellowship training. The finalized set of EPAs developed could become a foundation upon which standardized assessment, curriculum development, and program evaluation will be built.

## Data Availability

All data produced in the present study are available upon reasonable request to the authors.

## Contributions

Dr. Roumelioti served as the principal investigator and led all aspects of the study. She organized and facilitated the research procedures, submitted the IRB application, oversaw the development and dissemination of the surveys, and supervised the timely acquisition and accurate entry of survey data.

Dr. Singh co-supervised the study and contributed to all phases of the research. She facilitated participant recruitment, provided consultation over the development of the EPAs and the assessment tool, supported data collection and entry, and assisted in the dissemination of results.

Dr. Teixeira played a key role in recruitment and provided essential guidance on the development and evaluation of the EPAs. He also contributed to the creation of online surveys and the preparation and submission of abstracts and manuscripts for publication.

Dr. Argyropoulos provided oversight throughout the study and performed the statistical analyses necessary for data interpretation. He also contributed to the preparation of abstracts and manuscripts for dissemination of the study findings.

Dr. Isreal served as a consultant on the development and evaluation of the EPAs and contributed to overall study oversight.

D. Combs served as a reviewer and consultant for the preparation of this manuscript

Huzefa Y. Saria, second-year UNM medical student, was responsible for drafting and writing the manuscript and assisted in aligning developed EPAs with ACGME program accreditation milestones and sub competencies.

## Supplemental Tables

**Supplemental Table S1.** Guidelines for EPA Descriptions (Reproduced from Cate^6^)

|  |  |
| --- | --- |
| **Title** | Short, no words related to proficiency or skill.<br>Can a trainee be scheduled to do this? Can an entrustment decision for unsupervised practice for this EPA be made and documented? |
| **Description** | To enhance universal clarity: what is included?<br>What limitations apply? We will limit the description to the actual activity. Avoid saying why the EPA is important, or references to knowledge and skills. |
| **Required Knowledge, Skills, and Attitudes (KSAs):** | Which domains apply? Which sub-competencies apply? Include only the most relevant ones.<br>These links may serve to build observation and assessment methods. |
| **Required KSAs** | Which are necessary to execute the EPA? We will formulate this in a way to set expectations.<br>We plan to refer to resources that reflect necessary or helpful standards (books, a skills source, etc.) |
| **Information to assess progress** | We will consider observations, products, monitoring of knowledge skills, multi-source feedback. |
| **When is unsupervised practice expected** | Estimate acknowledging the flexible nature of this. |
| **Basis for formal entrustment decisions** | How many times must the EPA be executed proficiently for unsupervised practice? Who will judge this? What does formal entrustment look like? |

**Supplemental Table S2.**
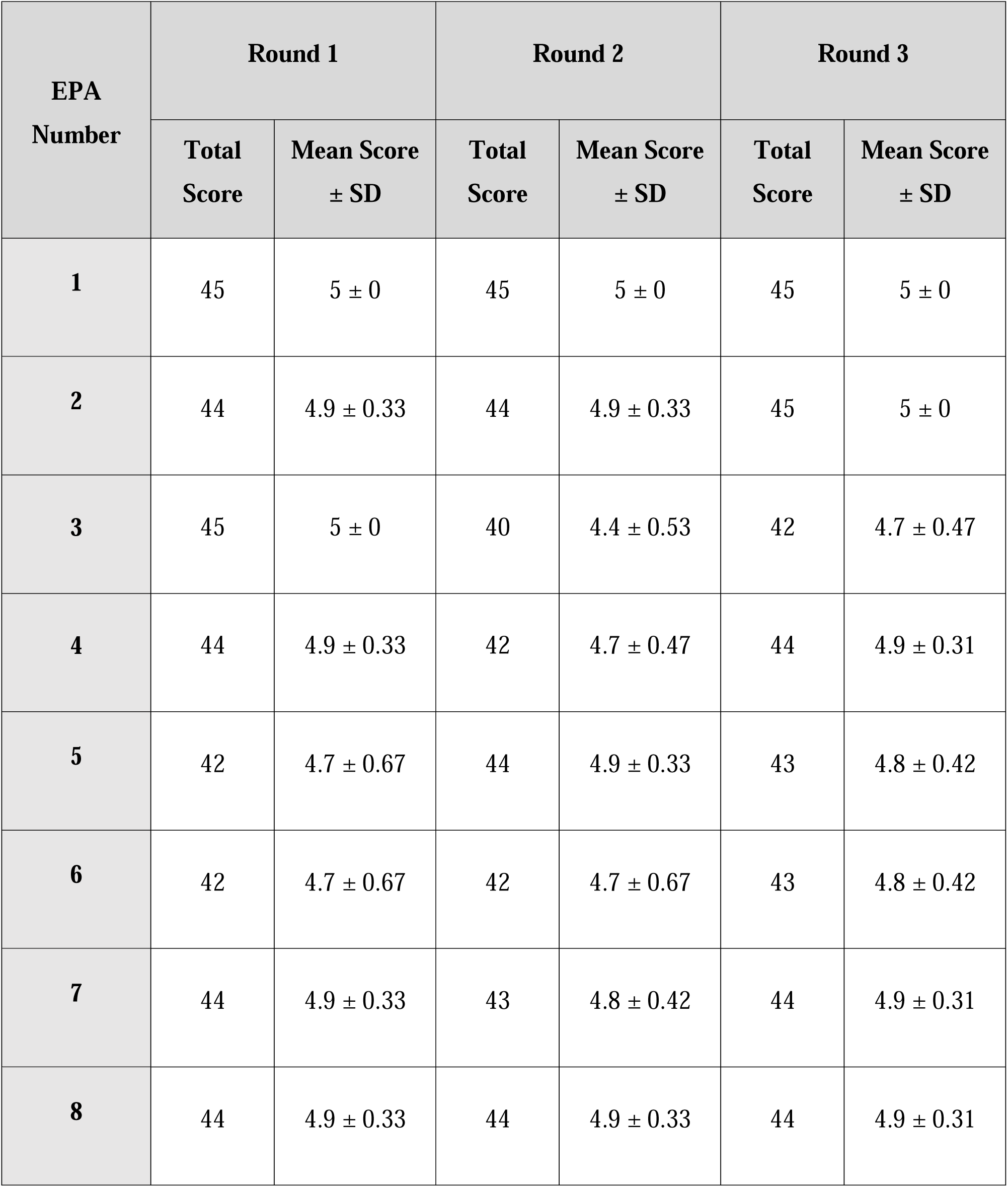
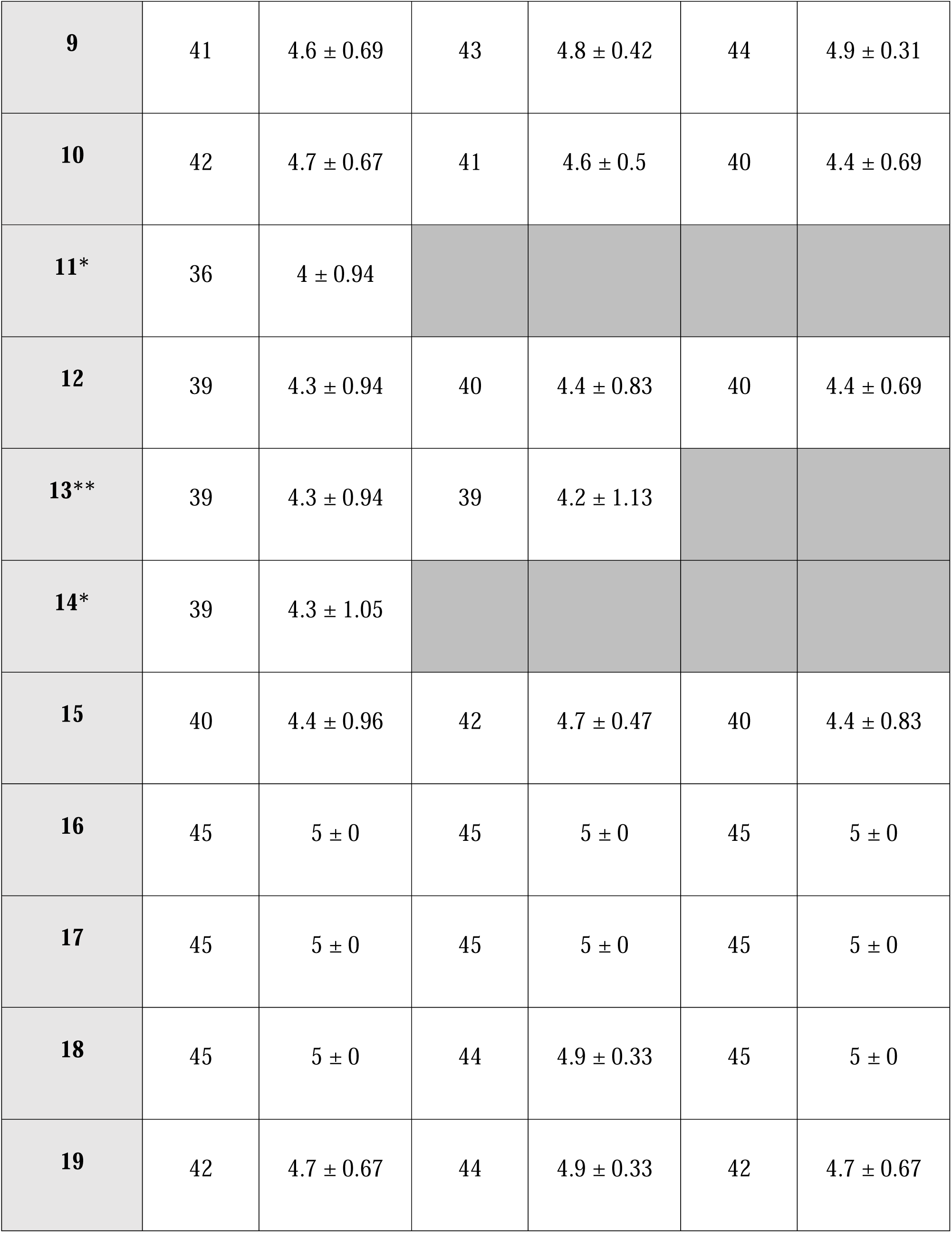
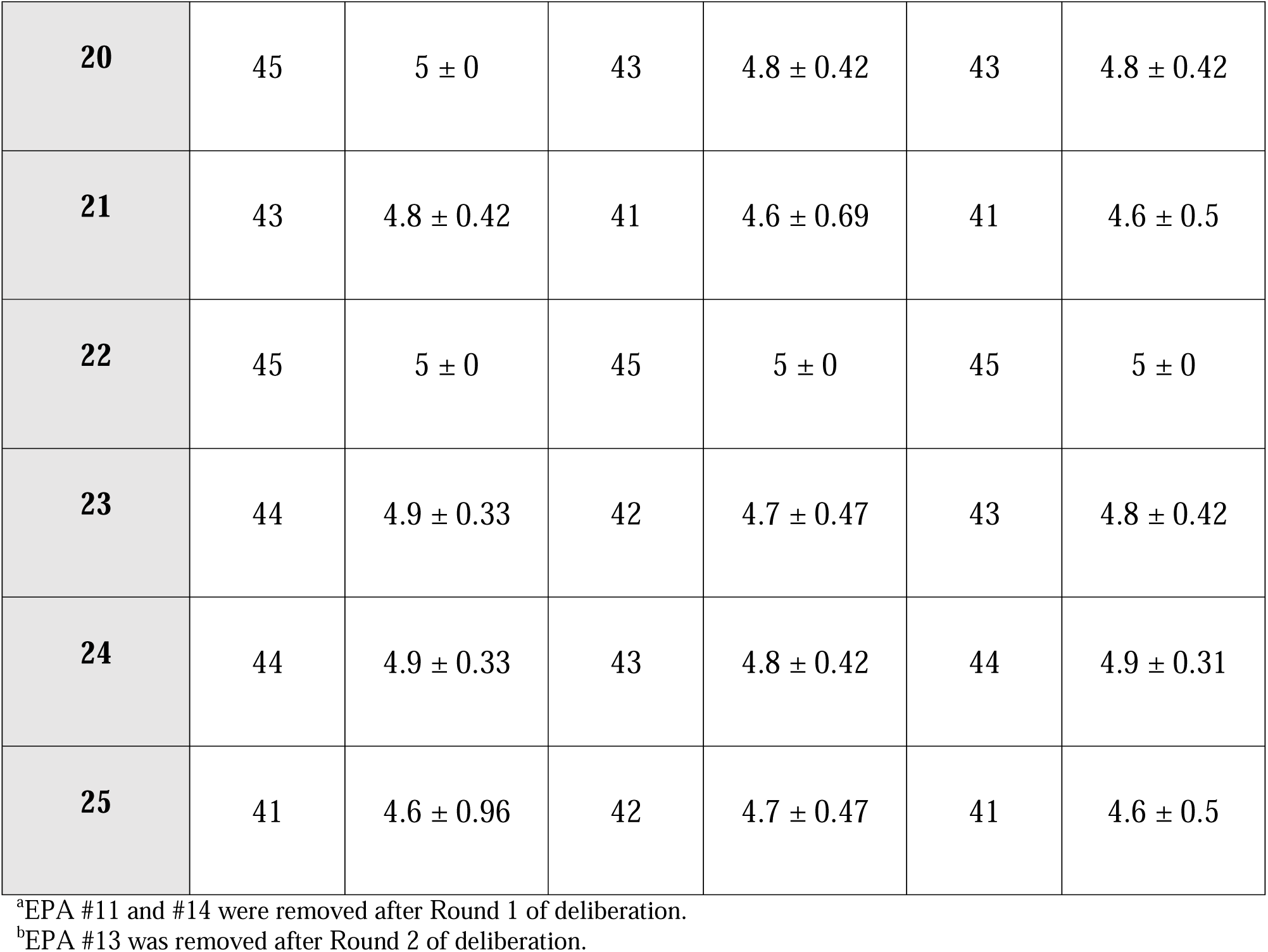
Round 1, 2, and 3 Questionnaire Results.

## Notes

### Competing Interest Statement

The authors have declared no competing interest.

### Funding Statement

This study was funded by the the Scholarship in Education Allocations Committee (SEAC)on 07/26/2023.
The SEAC Program is supported through funding from the Dean and administered by Continuous
Professional Learning and is is designed to stimulate and enhance the scholarship of education at the University of New Mexico School of Medicine

### Author Declarations

This study was approved by the Human Research Protection Program of the University of new Mexico Health Sciences Center (UNM-HSC) and granted approval on 9/28/2023 (Study ID 23-338)

## References

1. ten Cate O. Competency-Based Postgraduate Medical Education: Past, Present and Future. GMS J Med Educ. 2017;34(5):Doc69. doi:10.3205/zma001146

2. Touchie C, ten Cate O. The promise, perils, problems and progress of competency-based medical education. Med Educ. 2016;50(1):93–100. doi:10.1111/medu.12839

3. Cate O ten. A primer on entrustable professional activities. Korean J Med Educ. 2018;30(1):1–10. doi:10.3946/kjme.2018.76

4. Tanaka A, Kondo T, Urushibara-Miyachi Y, Maruyama S, Nishigori H. Development of entrustable professional activities for residents rotating nephrology department in a Japanese university hospital: a Delphi study. BMJ Open. 2021;11(8):e047923. doi:10.1136/bmjopen-2020-047923

5. Ten Cate O. Entrustability of professional activities and competency-based training. Med Educ. 2005;39(12):1176–1177. doi:10.1111/j.1365-2929.2005.02341.x

6. ten Cate O. Nuts and Bolts of table Professional Activities. J Grad Med Educ. 2013;5(1):157–158. doi:10.4300/JGME-D-12-00380.1

7. ACGME Nephrology Milestones. Accessed June 21, 2025. https://www.acgme.org/globalassets/pdfs/milestones/nephrologymilestones.pdf

8. Milestones by Specialty. Accessed June 21, 2025. https://www.acgme.org/milestones/milestones-by-specialty/

9. Torralba KD, Jose D, Katz JD. Competency-based medical education for the clinician-educator: the coming of Milestones version 2. Clin Rheumatol. 2020;39(6):1719–1723. doi:10.1007/s10067-020-04942-7

10. Yuan CM, Nee R, Abbott KC, Oliver JD. Milestones for Nephrology Training Programs: A Modest Proposal. Am J Kidney Dis. 2013;62(6):1034–1038. doi:10.1053/j.ajkd.2013.06.022

11. Yuan CM, Prince LK, Oliver JD, Abbott KC, Nee R. Implementation of Nephrology Subspecialty Curricular Milestones. Am J Kidney Dis. 2015;66(1):15–22. doi:10.1053/j.ajkd.2015.01.020

12. Pinsk M, Karpinski J, Carlisle E. Introduction of Competence by Design to Canadian Nephrology Postgraduate Training. Can J Kidney Health Dis. 2018;5:2054358118786972. doi:10.1177/2054358118786972

13. ACGME Program Requirements for Graduate Medical Education in Nephrology. Accessed August 3, 2025. https://www.acgme.org/globalassets/pfassets/programrequirements/2024-prs/148_nephrology_2024.pdf

